# Fusion Detection in Microsecretory Adenocarcinoma and Mucoepidermoid Carcinoma Using Chromogenic RNA In Situ Hybridization: A Promising Alternative to DNA-Based Fluorescence In-Situ Hybridization

**DOI:** 10.1101/2022.10.22.22281354

**Authors:** Doreen N. Palsgrove, Calvin Hosler, Lisa M. Rooper, Dequan Weston, Andrew Day, Justin A. Bishop, Richard C. Wang

## Abstract

**Background:** Recent advances in molecular genetics have dramatically improved our understanding of the pathophysiology and classification of salivary gland tumors. The identification of recurrent oncogenic fusions has been especially helpful in distinguishing entities with overlapping histomorphology.

**Methods:** Chromogenic RNA in situ hybridization (RNA-ISH) using BaseScope™ technology was performed to detect gene fusions associated with microsecretory adenocarcinoma (MSA), *MEF2C*::*SS18*, and mucoepidermoid carcinoma (MEC), *CRTC1*::*MAML2*, using probes specific to the exon junctions of the *MEF2C*::*SS18* (exon 7 of *MEF2C* to exon 4 of *SS18*) and *CRTC1*::*MAML2* (exon 1 of *CRTC1* to exon 2 of *MAML2*) fusion transcripts. Sixteen cases of *MEF2C*::*SS18* fusion-positive MSA, six cases of *CRTC1*::*MAML2* fusion-positive MEC, three cases of fusion-unknown MEC, and one case of fusion-negative MEC were included in the test cohort. Positive signal strength was assessed using a semi-quantitative scoring method as per manufacturer guidelines.

**Results:** Fusion transcripts were detected by RNA-ISH results in 14/16 cases (88%) of fusion-positive MSAs and 3/6 cases (50%) of fusion-positive MEC. Interestingly, 2 cases (67%) of fusion-unknown MEC were also positive by RNA-ISH for *CRTC1*::*MAML2* while the fusion-negative MEC was also negative by RNA-ISH. Positivity ranged between 1+ (one dot per cell in ≥5% of tumor cells in one 40X field) and 2+ (two to three dots per cell in ≥5% of tumor cells in one 40X field).

**Conclusion:** Here, we provide the first assessment of chromogenic RNA-ISH to detect gene fusions associated with microsecretory adenocarcinoma, *MEF2C*::*SS18*, and mucoepidermoid carcinoma, *CRTC1*::*MAML2*. Our results highlight the potential for ultrasensitive RNA-ISH to be used as an alternative method of fusion detection for salivary gland malignancies with highly conserved fusion transcript exon junctions. While additional studies are needed to validate the clinical utility of the assay and to determine optimal testing conditions, RNA-ISH may provide a means for restricted fusion analysis in cases with limited material and for pathologists without easy access to conventional molecular diagnostic testing.

## INTRODUCTION

The diagnosis of tumors continues to be based on multiple clinicopathologic parameters but with refined classification criteria predicated on an integrated diagnostic approach utilizing morphologic, immunophenotypic, and molecular data. Most oncologic processes involve gene fusions, rearrangements, and/or mutations and many tumor types, particularly newly recognized entities, are defined by their genetic abnormalities where possible.

Many salivary gland neoplasms harbor tumor-specific rearrangements, including both well-recognized and newly described entities. Mucoepidermoid carcinoma (MEC) is the most common salivary malignancy in all age groups, representing approximately a third of all malignant salivary gland tumors, molecularly characterized by fusions involving *MAML2* (1). The majority of MECs harbor t(11;19)(q21;p13), resulting in a canonical in-frame *CRTC1*::*MAML2* fusion, with a small subset having *CRTC3*::*MAML2* fusion secondary to translocation t(11;15)(q21;q26). *CRTC1/3*::*MAML2* gene fusions have been reported in approximately 78.4% of MECs using RT-PCR, with a *CRTC3* fusion partner occurring in about 2% of cases (2). Although most MECs can be diagnosed based on histologic features alone, variant morphologies including MEC without immunohistochemical evidence of squamoid cells, can present diagnostic challenges that would require molecular studies to resolve (3-5). Microsecretory adenocarcinoma (MSA) is a relatively new salivary gland entity with highly consistent histologic and immunophenotypic features (S100 and p63 positivity with negative p40) and recurrent *MEF2C*::*SS18* fusions involving exon 7 of *MEF2C* (5q14.3) and exon 4 of *SS18* (18q11.2). Like MEC, the presence of a *MEF2C*::*SS18* fusion is considered a useful diagnostic adjunct to distinguish it from other intermediate grade salivary carcinomas (6).

Molecular testing of salivary gland tumors is often performed for differential diagnostic accuracy and appropriate clinical management with fluorescence in situ hybridization (FISH) assays being the most commonly used method of detection for rearrangement, fusion, or aneuploidy of locus specific DNA sequences. FISH is considered a standard technique in many clinical molecular laboratories with the advantage of direct spatial localization of genetic abnormalities within tumor cells. Unfortunately, FISH is time consuming, requires the use of fluorescent probes (fluorescence gradually weakens over time), and can only be analyzed using specialized equipment (epifluorescence microscope with an adjusted set of filters) (7). Other less commonly used non-in situ methods include RT-PCR (reverse transcriptase-polymerase chain reaction) and NGS (next-generation sequencing). RT-PCR and NGS both face challenges related to the fragmentation of RNA in formalin-fixed paraffin-embedded (FFPE) tissue samples, and NGS has the additional barriers of higher cost, a longer workflow, and complex analysis (7).

An alternative approach for in situ testing is the use of chromogenic RNA in situ hybridization (RNA-ISH) (8). This technique is frequently used to determine the presence of high-risk human papilloma virus (HPV), which is required for the diagnosis of certain head and neck tumors (e.g., HPV-related multiphenotypic sinonasal carcinoma) (9, 10), and Epstein-Barr Virus (EBV) for primary nasopharyngeal carcinomas. Chromogenic RNA-ISH allows for the visualization of a specific RNA locus with spatial and morphological context and can be analyzed using a standard brightfield microscope. More recently, highly sensitive chromogenic RNA-ISH protocols (e.g., BaseScope™, Advanced Cell Diagnostics, Inc.) have been developed to enable detection of short RNA targets, circular RNA, and exon junctions/splice variants, and even point mutations (11-18).

Herein, we show that BaseScope™ RNA-ISH has the potential to be used as a clinical assay for the identification of fusion transcripts in salivary gland tumors such as MSA and MEC.

## METHODS

### Cases

Twenty cases of microsecretory adenocarcinoma (MSA) with available material were identified from consultation files (2015 to 2021). The presence of the *MEF2C*::*SS18* fusion – all fusion breakpoints involved exon 7 of the *MEF2C* gene (NCBI Reference Sequence: NM_002397.4) and exon 4 of *SS18* gene (NM_001007559.2) – was previously demonstrated by targeted RNA sequencing (RNA-seq) and/or RT-PCR in 18 of 20 cases of MSA, while two cases harbored *SS18* fusions with alternate gene partners (19, 20). Only archived unstained slides (stored at room temperature in standard archival conditions without a desiccant) were available for all cases of MSA.

Twelve cases of mucoepidermoid carcinoma (MEC) with available material were identified from consultation and institutional files (2017 and 2022). The presence of the *CRTC1*::*MAML2* fusion was previously demonstrated by targeted RNA-seq in 7 of 12 cases of MEC, while four cases of MEC had no molecular data, one case was negative for *MAML2* rearrangement by FISH, and one case was negative for fusions by RNA-seq. Only archived unstained slides (stored at room temperature in standard archival conditions without a desiccant) were available in 11 cases of MEC, while one case had freshly cut slides made from an archived formalin-fixed paraffin-embedded (FFPE) tissue block (stored at room temperature in standard archival conditions without a desiccant).

### BaseScope™ RNA in situ hybridization (ISH) ssay

The BaseScope™ Reagent Kit v2-RED (catalog no. 323910; Advanced Cell Diagnostics, Inc., Newark, CA, USA) was used to manually perform the BaseScope™ RED assay on FFPE tissue sections of MSA and MEC per manufacturer guidelines. Custom RNA probes (Advanced Cell Diagnostics, Inc.) were designed to target fusion transcript-specific exon junctions of interest (Figure 1) for MSA (*MEF2C*::*SS18*) and MEC (*CRTC1*::*MAML2*) based on known exon junctions reported in the literature (19-21) (BaseScope™ Probes, BA-Hs-MEF2C-SS18-fusion-Junc-C1, catalog no. 1195231-C1; BA-Hs-CRTC1-MAML2-fusion-Junc-C1, catalog no. 1195241-C1). Unstained FFPE 5-micron thick tissue sections were baked in a HybEZ™ (Advanced Cell Diagnostics, Inc.) oven at 60°C for 1 hour, followed by a series of deparaffinization steps. The sections were then left to dry at 60°C for 5 minutes. Once dry, RNAscope® Hydrogen Peroxide (Advanced Cell Diagnostics, Inc.) was applied to each slide at room temperature and left to incubate for 10 minutes. The slides were then washed in distilled water twice. This step was followed by manual target retrieval in which 700 mL of 1x RNAscope® Target Retrieval Reagent was poured into a beaker on a hot plate. Once the solution was warmed to 100°C, the slides were left to incubate in solution for 15 minutes. The slides were quickly transferred into distilled water, washed in 100% ethanol, and left out to dry at room temperature. A hydrophobic barrier was then drawn around the tissue of interest using an ImmEdge™ hydrophobic barrier PAP pen (H-4000). The slides were placed inside the EZ-Batch™ Slide Rack (Advanced Cell Diagnostics, Inc.), where RNAscope® Protease III was added to each sample. The slide rack was then inserted in the HybEZTM humidity control tray and incubated in the HybEZ™ oven (40°C, 30 min), followed by one wash in distilled water. The MEC and MSA probes were placed in the oven in conjunction with the slide rack to equilibrate prior to use. Excess liquid was then removed from the slides post-incubation, probe mix was added and hybridized (oven at 40°C, 2 hrs), and slides were washed (1x RNAscope® Wash Buffer twice, 2 min) and stored overnight (room temperature, 5x saline sodium citrate (SSC) buffer). The following day, a series of 8 amplification steps was performed per manufacturer guidelines. RED working solution (1:60, BaseScope™ Fast Red-B to Fast Red-A) was applied (incubation for 10 min, room temperature) and slides were counterstained using 50% Gill’s Hematoxylin I staining solution (catalog no. HXGHE1PT; American MasterTech Scientific) (room temperature, 2 min). Samples were then moved to a staining dish containing tap water (repeated until the slides were clear and sections remained purple), dipped in 0.02% ammonia water (221228; Sigma-Aldrich), washed with tap water, left out to dry (60°C, 15 min), and mounted using VectaMount (64742-48-9; Vector Laboratories).

**Figure 1.**
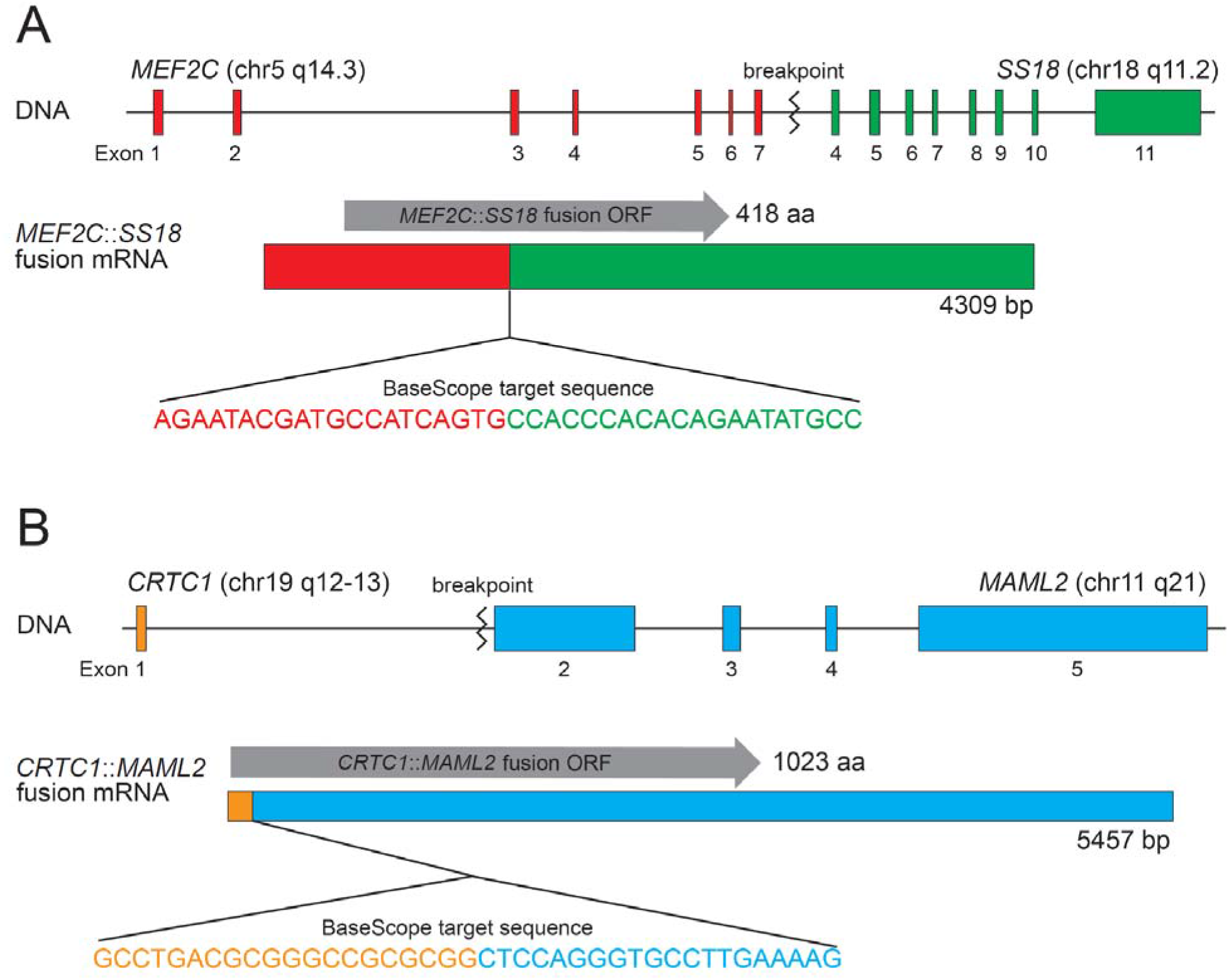
Diagrammatic representation of fusion transcript exon junctions of *MEF2C*::*SS18* (A) and *CRTC1*::*MAML2* (B).

The assay was performed separately for cases of MSA and MEC. Sequencing results were unblinded for one case each of MSA and MEC to serve as positive controls for tumor-specific fusion probes targeting the exon junctions of *MEF2C*::*SS18* and *CRTC1*::*MAML2*, respectively. Technical controls were performed on FFPE cultured cell pellets of human HeLa cells using a human-specific housekeeping gene positive control probe a non-specific bacterial (BaseScope™ Probes -BA-Hs-PPIB-1zz -Homo sapiens peptidylprolyl isomerase B (cyclophilin B) (PPIB) mRNA, catalog no. 710171; BaseScope™ Duplex Negative Control Probe-DapB-1ZZ, catalog no. 700141). Positive and negative control probes were additionally performed on cases of MEC. Adjacent benign tissue was used as negative internal control.

### Interpretation of RNA in situ hybridization (ISH)

Interpretation was performed using a standard bright field microscope at 20-40X magnification. No prior knowledge of molecular status was known when scoring. Positive signal strength in tumor cells was assessed using a semi-quantitative scoring method as per manufacturer guidelines (Table 1): staining score 0 – no staining or less than one dot to every 20 cells (<5%); staining score 1 – one dot per cell in ≥5% of tumor cells in one 40X field; staining score 2 – two to three dots per cell in ≥5% of tumor cells in one 40X field; staining score 3 – four to ten dots per cell in ≥5% of tumor cells in one 40X field. Only distinct, punctate dots of red chromogen precipitate within each cell boundary were counted as positive signals. Fewer than one dot per 20 cells (<5%) was considered acceptable background staining in negative control tissue. Hotspot areas of signal within the tumor were used for final interpretation in large tissue sections. Cases with scores□ ≥□1 were identified as positive.

**Table 1.**
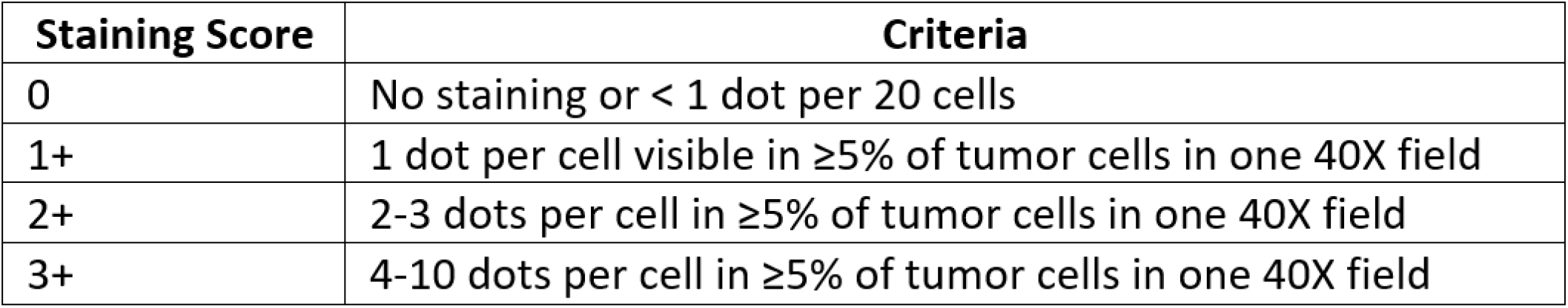
Semi-quantitative scoring adapted from manufacturer guidelines

| **Staining Score** | **Criteria** |
| --- | --- |
| 0 | No staining or < 1 dot per 20 cells |
| 1+ | 1 dot per cell visible in $\geq 5\%$ of tumor cells in one 40X field |
| 2+ | 2-3 dots per cell in $\geq 5\%$ of tumor cells in one 40X field |
| 3+ | 4-10 dots per cell in $\geq 5\%$ of tumor cells in one 40X field |

## RESULTS

### Microsecretory adenocarcinoma (MSA)

Sixteen of 20 cases of MSA survived processing and assay testing. Of the 16 cases evaluated, all were previously shown to harbor the *MEF2C*::*SS18* fusion (Table 2). 88% (14/16) of cases were considered positive using the described semi-quantitative scoring method – 7 cases demonstrated 1+ staining and 7 cases demonstrated 2+ staining (Figure 2). The two cases that were interpreted as negative showed rare punctate signals in scattered tumor cells, but these areas did not reach the 5% threshold to be considered positive.

**Table 2.** Microsecretory adenocarcinoma Basescope™ results

| Case No. | Procedure Year | Non-Tumor Score† | Tumor Score† | Overall Interpretation | Molecular |
| --- | --- | --- | --- | --- | --- |
| **MSA-1** | 2021 | 0 | 1 | + | *MEF2C::SS18* |
| **MSA-2\*** | 2020 | 0 | 0* | Negative* | *MEF2C::SS18* |
| **MSA-3** | 2020 | 0 | 2 | + | *MEF2C::SS18* |
| **MSA-4** | N/A | 0 | 1 | + | *MEF2C::SS18* |
| **MSA-5** | 2020 | 0 | 2 | + | *MEF2C::SS18* |
| **MSA-6** | 2021 | 0 | 2 | + | *MEF2C::SS18* |
| **MSA-7** | 2019 | 0 | 2 | + | *MEF2C::SS18* |
| **MSA-8** | 2020 | 0 | 2 | + | *MEF2C::SS18* |
| **MSA-9** | 2018 | 0 | 1 | + | *MEF2C::SS18* |
| **MSA-10** | 2019 | 0 | 1 | + | *MEF2C::SS18* |
| **MSA-11** | N/A | 0 | 1 | + | *MEF2C::SS18* |
| **MSA-12** | 2018 | 0 | 2 | + | *MEF2C::SS18* |
| **MSA-13** | 2016 | 0 | 1 | + | *MEF2C::SS18* |
| **MSA-14** | 2020 | 0 | 2 | + | *MEF2C::SS18* |
| **MSA-15** | N/A | 0 | 1 | + | *MEF2C::SS18* |
| **MSA-16\*** | 2015 | 0 | 0* | Negative* | *MEF2C::SS18* |

†, semi-quantitative scoring method: 0 - no staining or less than one dot to every 20 cells, 1 - one dot per cell, 2 - two to three dots per cell; \*, rare signals present in tumor cells but below threshold criteria to call positive; N/A, not available

**Figure 2.**
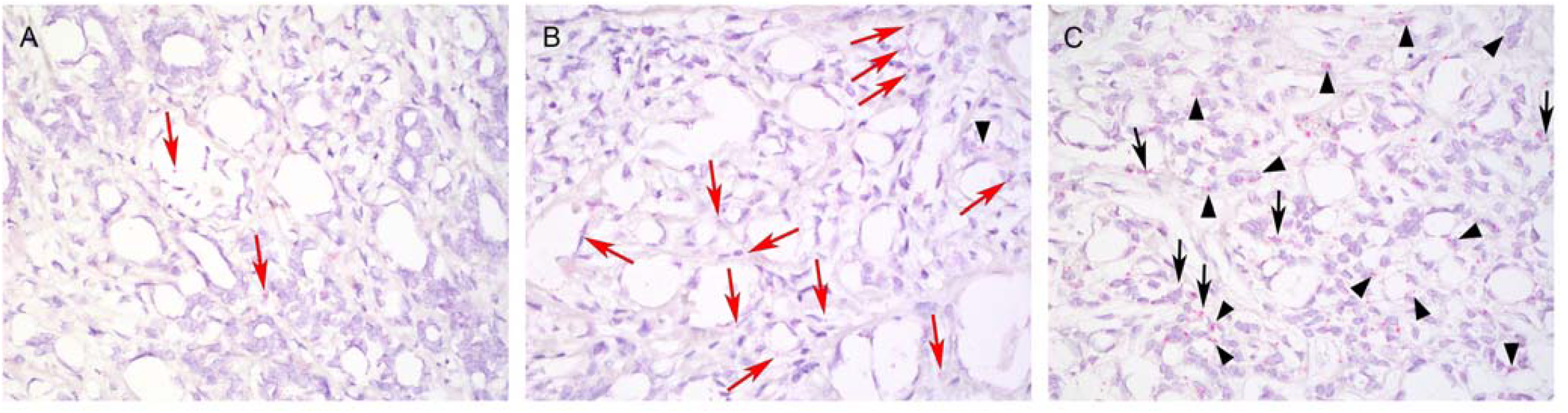
Representative images of *MEF2C*::*SS18* chromogenic RNA in situ hybridization in microsecretory adenocarcinoma. Positive signal strength was interpreted using a semi-quantitative scoring method as per manufacturer guidelines – 0, no staining or less than one dot to every 20 cells (A, MSA-2); 1+, one dot per cell in ≥5% of tumor cells in per 40X field (B, MSA-9); and 2+, two to three dots per cell in ≥5% of tumor cells in one 40X field (C, MSA-12). Red arrow – tumor cell with one dot; black arrowhead – tumor cell with 2 dots; black arrow – tumor cell with three dots.

### Mucoepidermoid carcinoma (MEC)

Ten of 12 cases of MEC survived processing and assay testing. Of the 10 cases evaluated, 6 cases were previously shown to harbor the *CRTC1*::*MAML2* fusion (Table 3), three had no molecular data available, and one was negative for *MAML2* rearrangement by break-apart FISH. Of the cases with known *CRTC1*::*MAML2* fusions, 50% (3/6) were considered positive (Figure 3). Two of 3 cases of MEC without available molecular data were also positive while one case showed rare signals in scattered tumor cells below threshold criteria to call positive.

**Table 3.** Mucoepidermoid carcinoma Basescope™ results

| Case No. | Procedure Year | Non-Tumor Score† | Tumor Score† | Overall Interpretation | Molecular |
| --- | --- | --- | --- | --- | --- |
| MEC-1* | 2021 | 0 | 0* | Negative* | N/A |
| MEC-2 | 2019 | 0 | 1 | + | N/A |
| MEC-3 | 2021 | 0 | 1 | + | *CRTC1::MAML2* |
| MEC-4 | 2022 | 0 | 0 | Negative | *CRTC1::MAML2* |
| MEC-5 | 2019 | 0 | 0 | Negative | *CRTC1::MAML2* |
| MEC-6 | N/A | 0 | 0 | Negative | *MAML2* FISH negative |
| MEC-7 | 2019 | 0 | 0 | Negative | *CRTC1::MAML2* |
| MEC-8 | 2021 | 0 | 1 | + | *CRTC1::MAML2* |
| MEC-9 | 2021 | 0 | 1 | + | N/A |
| MEC-10 | 2021 | 0 | 1 | + | *CRTC1::MAML2* |

†, semi-quantitative scoring method: 0 - no staining or less than one dot to every 20 cells, 1 - one dot per cell, 2 - two to three dots per cell; \*, rare signals present in tumor cells but below threshold criteria to call positive; N/A, not available

**Figure 3.**
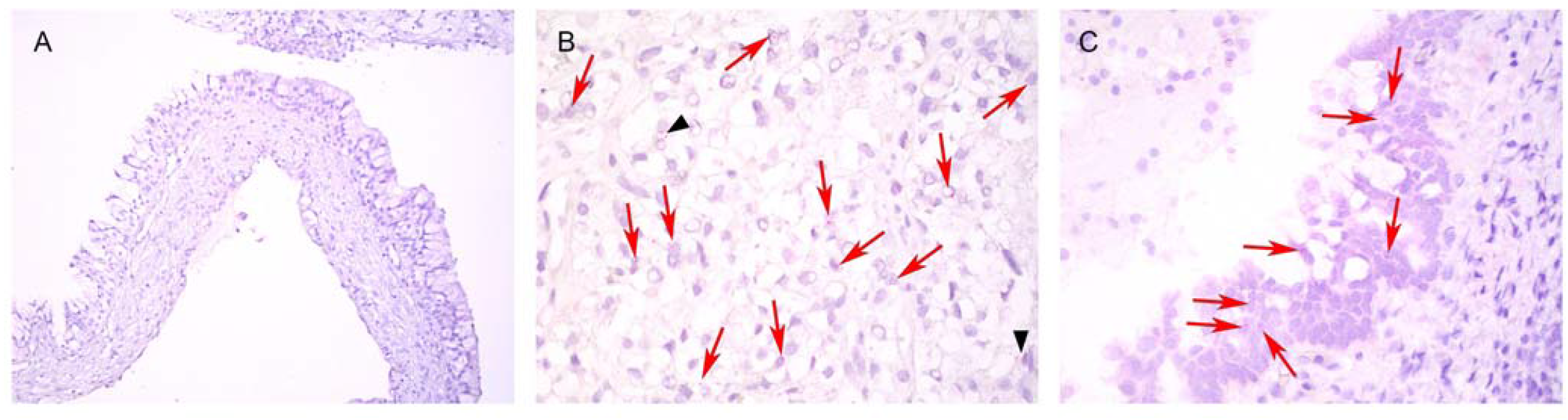
Representative images of *CRTC1*::*MAML2* chromogenic RNA in situ hybridization in mucoepidermoid carcinoma. Positive signal strength was interpreted using a semi-quantitative scoring method as per manufacturer guidelines – 0, no staining or less than one dot to every 20 cells (A, MEC-6) and 1+, one dot per cell in ≥5% of tumor cells in per 40X field (B, MEC-10; C, MEC-2). Red arrow – tumor cell with one dot; black arrowhead – tumor cell with 2 dots.

## DISCUSSION

In this study, chromogenic RNA-ISH was found to be a potentially useful diagnostic tool for the detection of *MEF2C*::*SS18* fusions in MSA and *CRTC1*::*MAML2* fusions in MEC. Fusion transcripts were detected by RNA-ISH results in 14/16 cases (88%) of fusion-positive MSAs and 3/6 cases (50%) of fusion-positive MEC. Interestingly, 2 cases (67%) of fusion-unknown MEC were also positive by RNA-ISH for *CRTC1*::*MAML2* while the fusion-negative MEC was also negative by RNA-ISH. Chromogenic BaseScope™ ISH was specific for both *MEF2C*::*SS18* and *CRTC1*::*MAML2* fusions in MSA and MEC, respectively. In addition, there was a high degree of concordance when comparing with RNA-Seq for MSA.

False-negative results are likely a consequence of RNA degradation and/or fusion transcripts expressed at low transcription levels (22). The BaseScope™ RNA targets are short (50 to 300 nucleotides) and assay performance depends on multiple preanalytical factors related RNA quality such as proper fixation (10% neutral buffered formalin for 16-32 hours, room temperature) and minimal tissue ischemia to reduce RNA degradation. In this study, archived unstained slides of various ages, stored at room temperature without a desiccant, were used in all but one case. Additionally, tissue sections for most cases were from large tumor resections where uniformity of tissue fixation may be less consistent compared to biopsy specimens. Fixation conditions and ischemic times were also unknown since most cases were obtained from outside contributors and these preanalytical variables are not routinely recorded for head and neck specimens.

In addition to chromogenic RNA-ISH, FISH and RT-PCR are other important adjunctive techniques for fusion detection. Both MSA and MEC can be molecular confirmed by commercially available break-apart FISH assays – *SS18* and *MAML2*, respectively – for definitive diagnosis in borderline cases; however, false-negatives are known to occur with both (6, 23, 24). Advantages of FISH over chromogenic RNA-ISH include 1) relative stability of DNA compared to RNA in archival FFPE tissue, and 2) the ability to assess for rearrangements using break-apart probes when break point are inconsistent and/or fusion partners are promiscuous. From a clinical application perspective, however, FISH assays cannot be used to identify small intrachromosomal rearrangements (e.g., *NCOA4*::*RET*), and cannot confirm that detected fusions are expressed (7). Resources required for implementation and interpretation, such as a fluorescence microscope and experienced staff for FISH studies, are also limited, making RNA-ISH an attractive option for pathology laboratories. Furthermore, RNA-ISH is adaptable to automated immunohistochemistry platforms, including Leica and Roche Ventana systems, allowing for more widespread adoption while minimizing hands-on time (25). Since the BaseScope™ assay utilizes the same platform of the established RNAscope™ technology its implementation for fusion detection in the clinical setting may become somewhat analogous to that of immunohistochemistry.

The notable limitations of RNA-ISH for fusion detection are similar to those of RT-PCR: 1) messenger RNA is inherently labile; 2) target sequences are limited to specific exon junctions of known gene partners; and 3), multiplexing capabilities are currently limited. In practice, RNA degradation is less of a concern since most tissues will have been newly obtained. Still, empiric thresholds of “positivity” need to be defined to ensure appropriate sensitivity and specificity, particularly in cases with minimal expression. To limit errors of interpretation due to signal degradation, tissue sections should be freshly cut from blocks that are less than one year old. Caution is also advised when implementing RNA-ISH for fusion detection on whole tissue sections as RNA preservation may be patchy and heterogeneous.

This study shows the feasibility of the BaseScope™ chromogenic RNA-ISH platform as a rapid diagnostic test for the detection of canonical fusion transcripts associated with MSA and MEC. Further studies are needed to fully assess the degree of sensitivity, specificity, and effectiveness of BaseScope™ as an independent fusion detection method.

## Data Availability

All data produced in the present work are contained in the manuscript

## Acknowledgements

Dr. Sukhbir Lulla (ACD Biosystems) is thanked for help in setting up the BaseScope assay.

## DECLARATIONS AND COMPLIANCE WITH ETHICAL STANDARDS

### Availability of data and material

not applicable.

### Code availability

not applicable.

### Ethics approval

All procedures performed in this retrospective data analysis involving human participants were in accordance with the ethical standards of the institutional review board (IRB 112017-073), which did not require informed consent. All authors confirm they have meaningfully contributed to the research and read and approved the final manuscript.

### Consent to participate

Waived by the IRB due to the retrospective nature of the work without therapeutic alterations.

### Consent for publication

Obtained from all individual participants for whom identifying information is uniquely included in this manuscript.

